# Rates of new indication for antithrombotic drugs in people with cognitive impairment: implications for anti-amyloid monoclonal antibody treatment

**DOI:** 10.1101/2025.07.17.25331746

**Authors:** Anna L Parks, Jacquelyn M Lykken, Meghan L Rieu-Werden, Darae Ko, Dae Hyun Kim, Margaret C Fang, Steven M Greenberg, Daniel M Witt, Mark A Supiano, Sachin J Shah

## Abstract

**Objectives:** Antithrombotic drugs—anticoagulants and thrombolytics— may interact with anti-amyloid monoclonal antibodies (mAbs) to increase intracranial hemorrhage risk, so expert guidance recommends against their co-prescription. We aimed to estimate how many people with mild cognitive impairment (MCI) or dementia develop a new cardiovascular indication for antithrombotic drugs.

**Methods:** In a longitudinal cohort of adults ≥65 years old from the Health and Retirement Study (2010-2020) with linked Medicare claims and no prior indication for anticoagulants, cognition was categorized as normal, MCI, or dementia. We fit Fine-Gray separate survival models accounting for competing risk of death to estimate 1-year incidence of atrial fibrillation (AF), deep vein thrombosis (DVT), pulmonary embolism (PE), acute myocardial infarction (MI), and stroke.

**Results:** Among 12,373 participants (mean age 73 years, 59% female), for MCI, 1-year incidence was 1.7% for AF, 1.2% for DVT, 0.4% for PE, 1.2% for AMI, 2.0% for stroke, and 5.7% for any indication. In dementia, 1-year rates were 1.7% for AF, 1.8% for DVT, 0.3% for PE, 1.0% for AMI, 2.4% for stroke, and 6.7% for any indication.

**Discussion:** Our finding inform shared decision-making about the tradeoffs of anti-amyloid mAbs.

## Introduction

The anti-amyloid monoclonal antibodies (mAbs) lecanemab and donanemab modestly slow cognitive decline in people with mild cognitive impairment (MCI) or mild dementia due to Alzheimer’s disease. The major adverse effects of this medication class are amyloid-related imaging abnormalities (ARIA), which rarely can present as intracranial hemorrhage (ICH). Appropriate use recommendations advise against co-prescribing antithrombotic drugs—including anticoagulants and thrombolytics—due to potential interactions with anti-amyloid mAbs that increase ICH risk.^1,2^

Because new cardiovascular diagnoses, such as atrial fibrillation (AF) or pulmonary embolism (PE), are common in older adults, some people may develop an indication for antithrombotic drugs during an anti-amyloid mAb treatment course.^3^ Professional societies, appropriate use recommendations and expert guidance recommend shared decision-making for anti-amyloid mAb treatment decisions.^4–8^ These discussions require individualized estimates of risks and benefits. Understanding the risk of developing a new condition where antithrombotic drugs are indicated during anti-amyloid mAb therapy is important when considering the tradeoffs of treatment. Our goal was to estimate how many people with MCI or dementia develop a new condition for which anticoagulants or thrombolytics are indicated.

## Methods

### Participants

We conducted a longitudinal cohort study using data from 2010-2020 from the Health and Retirement Study (HRS). The HRS is a nationally representative, longitudinal cohort of Americans over age 50 with linked Medicare claims. Subjects are interviewed biennially on topics including health and cognition. Proxies are interviewed when participants cannot answer questions due to physical or cognitive impairment. We included HRS participants aged 65 and older with Medicare claims linkage, at least 12 months of continuous Medicare Part A, B, and D enrollment, no diagnosis of AF or anticoagulant prescription in the 12 months prior to a baseline interview, and no missing information on cognition (**Supplementary Figure 1**). Participants were enrolled when they first met inclusion criteria and censored at disenrollment from Medicare, end of 12-month follow-up, or death.

### Covariates

To assess cognition, HRS researchers administer a validated 27-question neuropsychological battery to participants or an informant questionnaire to proxies when participants cannot be interviewed.^9^ We used standardized definitions that categorize participants’ cognition as normal, MCI, or dementia (defined in HRS as moderate or severe cognitive impairment), which correlate with clinically diagnosed MCI and dementia.^10^ We used claims-based algorithms (**Supplementary Table 1**) to define AF, deep vein thrombosis (DVT), PE, myocardial infarction (MI), and stroke.

### Statistical Analysis

After describing the cohort demographics, we fit separate Fine-Gray survival models accounting for competing risk of death to estimate the 1-year incidence of each outcome. Models used a robust sandwich variance estimator accounting for repeated observations.

### Standard Protocol Approvals, Registrations, and Participant Consents

The institutional review board at Mass General Brigham approved the study as exempt, and we followed STROBE reporting guidance.

### Data Availability

Researchers can apply to the Health and Retirement Study (https://hrs.isr.umich.edu/) to access the data used in this study.

## Results

The study included 12,373 participants (mean age 73 years, 59% female), including 2,433 (20%) with MCI, and 1,145 (9%) with dementia (**Table**). Among participants with MCI, the estimated 1-year incidence was 1.7% for AF, 1.2% for DVT, 0.4% for PE, 1.2% for MI, and 2.0% for stroke (**Figure**). The estimated 1-year incidence of developing a new indication for antithrombotic drugs for any reason was 5.7%.

**Figure 1:**
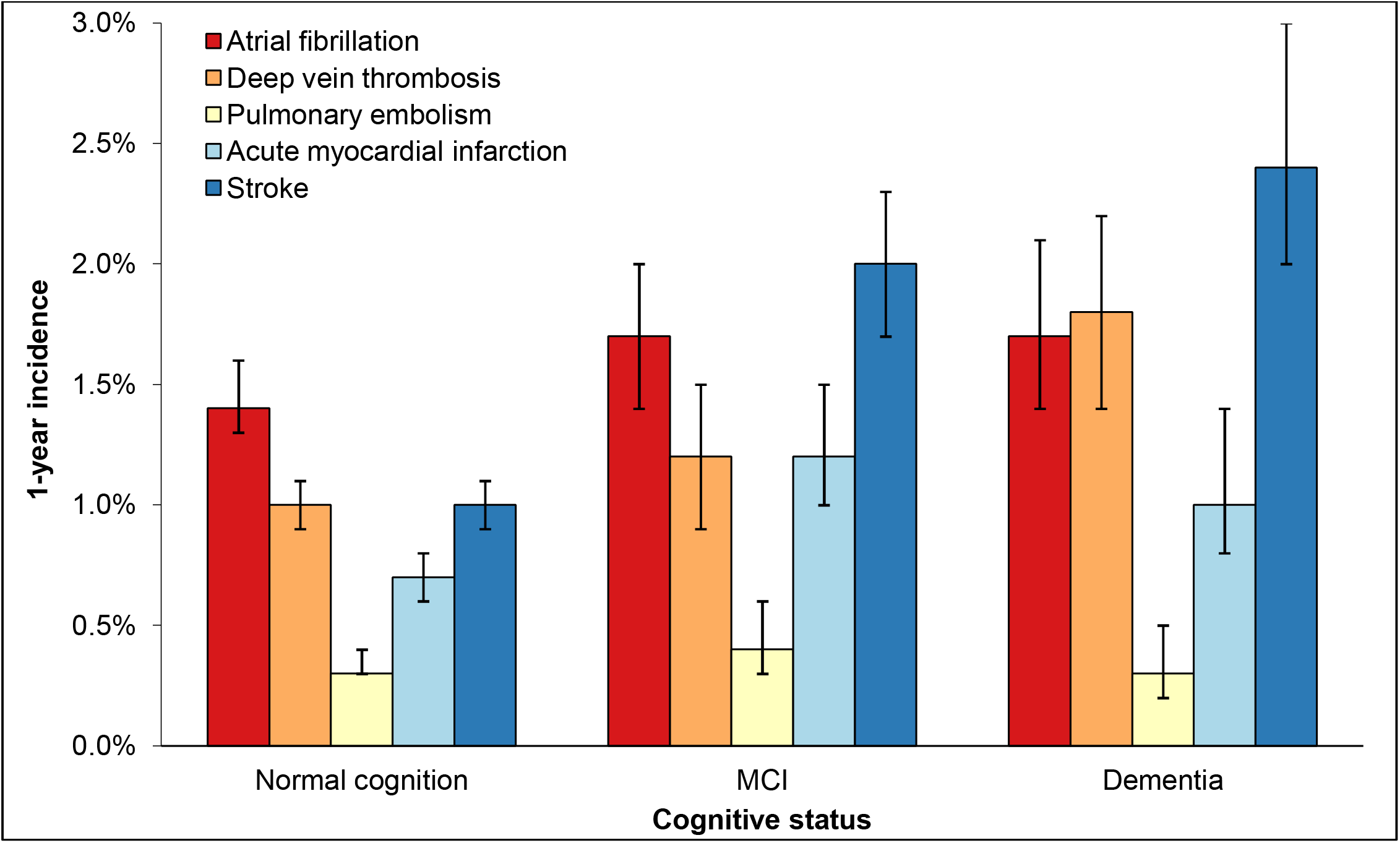
1-year incidence of cardiovascular disease by cognitive status.

**Table 1:** Cohort sociodemographics, geriatric syndromes, and comorbidities stratified by cognition.

**Table: Cohort sociodemographics, geriatric syndromes, and comorbidities stratified by cognition**
|  | No cognitive impairment | Mild cognitive impairment | Dementia |
| --- | --- | --- | --- |
| **Total Unique Cohort Members** | 8,795 | 2,433 | 1,145 |
| **Sociodemographics** |  |  |  |
| Age, mean (SD) | 72 (7) | 75 (8) | 80 (9) |
| Female | 59% | 56% | 63% |
| Race |  |  |  |
| Black/African American | 12% | 25% | 28% |
| White/Caucasian | 83% | 67% | 65% |
| Other* | 5% | 7% | 6% |
| Hispanic | 9% | 16% | 15% |
| High School graduate | 69% | 88% | 91% |
| Partnered/Married | 65% | 53% | 36% |
| Nursing home resident | 0.3% | 3% | 24% |
| Proxy interview | 2% | 6% | 46% |
| **Geriatric Syndromes\*\*** |  |  |  |
| Fall in the past 2 years | 33% | 39% | 54% |
| Injurious fall | 28% | 32% | 39% |
| Impaired vision | 19% | 37% | 46% |
| Impaired hearing | 20% | 33% | 41% |
| ADL dependence | 12% | 29% | 61% |
| IADL dependence | 10% | 28% | 68% |
| Incontinence | 27% | 27% | 39% |
| **Comorbidities\*\*\*** |  |  |  |
| Heart Failure | 11% | 22% | 33% |
| Prior MI | 8% | 11% | 14% |
| Angina | 4% | 5% | 4% |
| BMI |  |  |  |
| <18.5 | 1% | 2% | 8% |
| 18.5-24.9 | 28% | 32% | 41% |
| 25.0-29.9 | 39% | 36% | 30% |
| ≥30.0 | 33% | 30% | 22% |
| Prior hip fracture | 1% | 4% | 11% |
| Osteoporosis | 13% | 15% | 23% |
| Self-reported fair/poor health | 21% | 42% | 59% |
| Hypertension | 63% | 70% | 75% |
| Diabetes | 29% | 42% | 41% |
| Ever smoked tobacco | 56% | 58% | 52% |
| Prior stroke/TIA | 10% | 19% | 34% |
| Ischemic heart disease | 26% | 38% | 46% |
| Anemia<sup>3</sup> | 26% | 40% | 55% |
| Prior cancer diagnosis (except minor skin cancer) | 19% | 20% | 23% |
| Lung disease | 10% | 14% | 13% |
| Chronic kidney disease | 9% | 17% | 23% |

Cohort members stratified by cognitive status identified at first interview. Numbers reported as percent of non-missing (count / non-missing total).
Abbreviations: IQR=interquartile range, ADL=activities of daily living, IADL=instrumental activities of daily living, MI=myocardial infarction, TIA=transient ischemic attack
\* Other includes a pooled group of participants who identified as American Indian, Alaskan Native, Asian, Native Hawaiian, and Pacific Islander or another self-described race, consolidated due to small sample sizes and risk of identification.
\*\* Geriatric syndromes, angina, self-reported fair/poor health, tobacco use, and lung disease were derived from self-report in the Health and Retirement Study survey. Prior myocardial infarction, hip fracture, osteoporosis, hypertension, diabetes, prior stroke/TIA were derived from Medicare claims. Ischemic heart disease, heart failure, anemia, and chronic kidney disease were derived from a combination of self-report and Medicare claims.

Among participants with dementia, estimated 1-year incidence was 1.7% for AF, 1.8% for DVT, 0.3% for PE, 1.0% for MI, and 2.4% for stroke. Overall, in those with dementia, 6.7% developed a new indication for antithrombotic drugs for any reason in 1 year.

## Discussion

Over 1 year, 6-7% of people with MCI or dementia in this national sample were newly diagnosed with a cardiovascular disease where anticoagulants or thrombolytics are indicated. As clinicians, patients and caregivers consider anti-amyloid mAbs, these results should be added to the risk-benefit discussion.^11^

Other risk factors for ICH in patients on anti-amyloid therapy can be screened for and potentially mitigated either through patient selection (such as cerebral amyloid angiopathy and APOE-ε4 genotype) or treatment intensification (such as hypertension).^12^ In addition, clinicians can already use clinical trial and extension-phase data on ICH rates based on these fixed risk factors to inform discussion. In contrast, antithrombotic use is substantially more dynamic and detailed risk estimates have not been available until now.

Our results provide estimates of the likelihood of emergent cardiovascular conditions that can be incorporated into the decision to start anti-amyloid mAbs, as well as advance care planning. We found that the 1-year risk of 3 common acute clinical conditions—stroke, PE and MI—was 2.0-2.4%, 0.3-0.4% and 1.0-1.2%, respectively. These conditions at the very least require rapid decision-making and treatment with anticoagulants, and guidelines recommend administration of thrombolytics for patients with stroke who present within the appropriate timeframe, PE who have hemodynamic instability, and MI if percutaneous coronary intervention is not available. Due to the uncertainty and potential for catastrophic ICH, both lecanemab and donanemab appropriate use recommendations recommend against treating patients with anticoagulants or thrombolytics.^2,5^ Clinicians should discuss the small but potentially fatal or disabling possibility of patients not being able to receive emergent treatment for these acute conditions while receiving anti-amyloid mAbs so that patients and caregivers have a fully informed understanding of the tradeoffs.

Our results also inform discussion of the development of non-emergent conditions like a new diagnosis of AF (1.7% per year) or DVT (1.2-1.8%). Most older adults with AF and MCI or dementia will qualify for anticoagulation to prevent ischemic stroke based on their CHA_2_DS_2_-VAS_c_ score.^13^ And guideline-recommended therapy for DVT and PE is at least 3 months of anticoagulant treatment and long-term anticoagulation in those with thromboses that occur in the absence of provoking risk factors.^14^ Although recommendations about concurrent anticoagulant use may shift over time, most centers currently exclude these patients. Annual risk of stroke is 5-7% in AF without anticoagulation, and the annual risk of recurrent venous thromboembolism is between 3-10%. Thus, our results can be used to discuss with patients the possibility of having to decide between continuing anti-amyloid therapy versus forgoing anticoagulation for these chronic conditions.

Our data have limitations. We lack the granularity to determine eligibility for anti-amyloid therapy, and risk estimates may differ for people who qualify for anti-amyloid treatment. Additionally, while not all patients with new cardiovascular conditions would qualify for thrombolytic or anticoagulant therapy, a substantial proportion would meet guideline indications, which justifies consideration of this possibility.

The availability of anti-amyloid mAbs is a paradigm shift in dementia care. Incorporating the considerable risk of developing a condition for which antithrombotic drugs are indicated would enhance shared decision-making for these novel therapies.

## References

1. Cummings J, Apostolova L, Rabinovici GD, et al. Lecanemab: Appropriate Use Recommendations. J Prev Alzheimers Dis. 2023;10(3):362–377. doi:10.14283/jpad.2023.30

2. Rabinovici GD, Selkoe DJ, Schindler SE, et al. Donanemab: Appropriate use recommendations. J Prev Alzheimers Dis. 2025;12(5):100150. doi:10.1016/j.tjpad.2025.100150

3. Ko D, Pascual-Leone A, Shah SJ. Use of Lecanemab for Patients With Cardiovascular Disease: The Challenge of Uncertainty. JAMA. Published online March 15, 2024. doi:10.1001/jama.2024.2991

4. Ramanan VK, Armstrong MJ, Choudhury P, et al. Antiamyloid Monoclonal Antibody Therapy for Alzheimer Disease. Neurology. 2023;101(19):842–852. doi:10.1212/WNL.0000000000207757

5. Cummings J, Apostolova L, Rabinovici GD, et al. Lecanemab: Appropriate Use Recommendations. J Prev Alz Dis. Published online 2023. doi:10.14283/jpad.2023.30

6. Greenberg SM, Aparicio HJ, Furie KL, et al. Vascular Neurology Considerations for Antiamyloid Immunotherapy: A Science Advisory From the American Heart Association. Stroke. 2025;56(1):e30–e38. doi:10.1161/STR.0000000000000480

7. Sarkisian CA, Romanov A, Mafi JN. Talking With Patients About the New Anti-amyloid Alzheimer Disease Medications. Ann Intern Med. 2024;177(2):246–248. doi:10.7326/M23-3377

8. Monoclonal anti□amyloid antibodies for the treatment of Alzheimer’s disease and the hesitant geriatrician - Chin - 2024 - Journal of the American Geriatrics Society - Wiley Online Library. Accessed March 21, 2025. https://agsjournals.onlinelibrary.wiley.com/doi/10.1111/jgs.18652

9. Crimmins EM, Kim JK, Langa KM, Weir DR. Assessment of Cognition Using Surveys and Neuropsychological Assessment: The Health and Retirement Study and the Aging, Demographics, and Memory Study. J Gerontol B Psychol Sci Soc Sci. 2011;66B(Suppl 1): i162–i171. doi:10.1093/geronb/gbr048

10. Langa KM, Plassman BL, Wallace RB, et al. The Aging, Demographics, and Memory Study: study design and methods. Neuroepidemiology. 2005;25(4):181–191. doi:10.1159/000087448

11. Widera EW, Brangman SA, Chin NA. Ushering in a New Era of Alzheimer Disease Therapy. JAMA. 2023;330(6):503–504. doi:10.1001/jama.2023.11701

12. Zimmer JA, Ardayfio P, Wang H, et al. Amyloid-Related Imaging Abnormalities With Donanemab in Early Symptomatic Alzheimer Disease: Secondary Analysis of the TRAILBLAZER-ALZ and ALZ 2 Randomized Clinical Trials. JAMA Neurology. Published online March 10, 2025. doi:10.1001/jamaneurol.2025.0065

13. Joglar JA, Chung MK, Armbruster AL, et al. 2023 ACC/AHA/ACCP/HRS Guideline for the Diagnosis and Management of Atrial Fibrillation: A Report of the American College of Cardiology/American Heart Association Joint Committee on Clinical Practice Guidelines. Circulation. 2024;149(1):e1–e156. doi:10.1161/CIR.0000000000001193

14. Stevens SM, Woller SC, Kreuziger LB, et al. Antithrombotic Therapy for VTE Disease: Compendium and Review of CHEST Guidelines 2012-2021. CHEST. 2024;166(2):388–404. doi:10.1016/j.chest.2024.03.003

