## Supplement for "Rates of new indication for antithrombotic drugs in people with cognitive impairment: implications for anti-amyloid monoclonal antibody treatment"

**Table of contents:**

**Appendix Figure 1: Cohort flow diagram**

**Appendix Table 1: Cognition and outcome definitions**

**Appendix Figure 1: Cohort flow diagram**

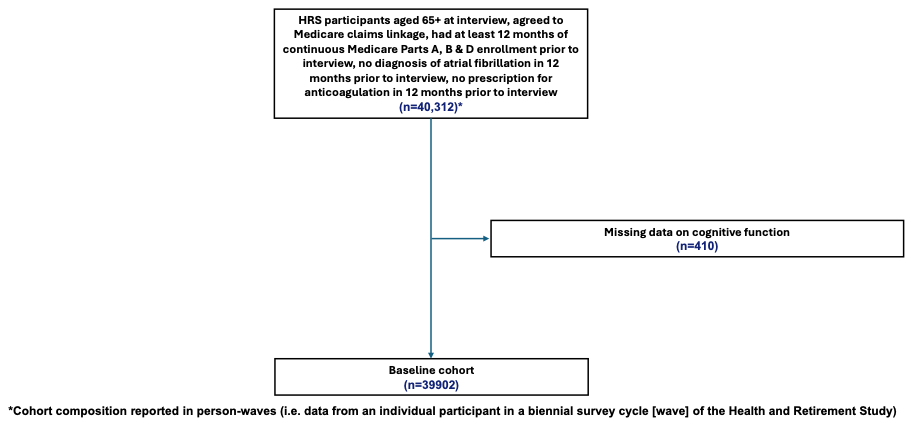

**Appendix Table 1: Cohort outcome definitions**

| **Variable** | **Criteria** | **Definition** |
| --- | --- | --- |
| *Cohort inclusion/Cognitive function*^3,4^ | | |
| Normal cognition | Respondents: Langa-Weir classification based on 27-point cognitive scale administered to respondents assessing immediate and delayed 10-noun free recall test, serial 7 subtraction test, and backward count from 20 test. Validated against clinical diagnosis.  Proxies: Langa-Weir classification based on 11-point scale assessing proxy’s assessment of the respondent’s memory (excellent to poor, 0-4); limitations in instrumental activities of daily living (0-5); and survey interviewer’s assessment of the respondent’s difficulty completing interview due to cognitive limitation (0-2). | Respondents: Score 12-27  Proxies: Score 0-2 |
| Mild cognitive impairment | Respondents: Langa-Weir classification based on 27-point cognitive scale administered to respondents assessing immediate and delayed 10-noun free recall test, serial 7 subtraction test, and backward count from 20 test. Validated against clinical diagnosis.  Proxies: Langa-Weir classification based on 11-point scale assessing proxy’s assessment of the respondent’s memory (excellent to poor, 0-4); limitations in instrumental activities of daily living (0-5); and survey interviewer’s assessment of the respondent’s difficulty completing interview due to cognitive limitation (0-2). | Respondents: Score 7-11  Proxies: Score 3-5 |
| Dementia | Respondents: Langa-Weir classification based on 27-point cognitive scale administered to respondents assessing immediate and delayed 10-noun free recall test, serial 7 subtraction test, and backward count from 20 test. Validated against clinical diagnosis.  Proxies: Langa-Weir classification based on 11-point scale assessing proxy’s assessment of the respondent’s memory (excellent to poor, 0-4); limitations in instrumental activities of daily living (0-5); and survey interviewer’s assessment of the respondent’s difficulty completing interview due to cognitive limitation (0-2). | Respondents: Score 0-6  Proxies: Score 6-11 |
| *Outcomes* | | |
| Atrial fibrillation | At least 1 inpatient claim OR 2 HOP/Carrier claims with DX codes | ICD9: 427.31 (ONLY first or second DX on the claim)  ICD10: I48.0, I48.1, I48.11, I48.19, I48.2, I48.20, I48.21, I48.91 (ONLY first or second DX on the claim) |
| Deep vein thrombosis | At least 1 inpatient OR HOP/Carrier claim with DX codes | ICD9:  453.2 453.40 453.41 453.42 451.11 451.19 451.81  453.82 453.83 453.84 453.85 453.86 453.87 451.83  ICD10:  I80.10 I80.11 I80.12 I80.13 I80.201 I80.202 I80.203 I80.209 I80.211 I80.212 I80.213 I80.219 I80.221 I80.222 I80.223 I80.229 I80.231 I80.232 I80.233 I80.239 I80.241 I80.242 I80.243 I80.249 I80.251 I80.252 I80.253 I80.259 I80.291 I80.292 I80.293 I80.299 I82.220 I82.401 I82.402 I82.403 I82.409 I82.411 I82.412 I82.413 I82.419 I82.421 I82.422 I82.423 I82.429 I82.431 I82.432 I82.433 I82.439 I82.441 I82.442 I82.443 I82.449 I82.451 I82.452 I82.453 I82.459 I82.461 I82.462 I82.463 I82.469 I82.491 I82.492 I82.493 I82.499 I82.4Y1 I82.4Y2 I82.4Y3 I82.4Y9 I82.4Z1 I82.4Z2 I82.4Z3 I82.4Z9  I80.10 I80.11 I80.12 I80.13 I80.201 I80.202 I80.203 I80.209 I80.211 I80.212 I80.213 I80.219 I80.221 I80.222 I80.223 I80.229 I80.231 I80.232 I80.233 I80.239 I80.241 I80.242 I80.243 I80.249 I80.251 I80.252 I80.253 I80.259 I80.291 I80.292 I80.293 I80.299 I82.220 I82.401 I82.402 I82.403 I82.409 I82.411 I82.412 I82.413 I82.419 I82.421 I82.422 I82.423 I82.429 I82.431 I82.432 I82.433 I82.439 I82.441 I82.442 I82.443 I82.449 I82.451 I82.452 I82.453 I82.459 I82.461 I82.462 I82.463 I82.469 I82.491 I82.492 I82.493 I82.499 I82.4Y1 I82.4Y2 I82.4Y3 I82.4Y9 I82.4Z1 I82.4Z2 I82.4Z3 I82.4Z9 |
| Pulmonary embolism | At least 1 inpatient claim with DX codes | ICD9: 415.11 415.13 415.19  ICD10: I26.02 I26.09 I26.92 I26.93 I26.94 I26.99 |
| Acute myocardial infarction | At least 1 inpatient claim with DX codes | ICD9: 410.01, 410.11, 410.21, 410.31, 410.41, 410.51, 410.61, 410.71, 410.81, 410.91 (ONLY first or second DX on the claim)  ICD10: I21.01, I21.02, I21.09, I21.11, I21.19, I21.21, I21.29, I21.3, I21.4, I21.9, I21.A1, I21.A9, I22.0, I22.1, I22.2, I22.8, I22.9 (ONLY first or second DX on the claim) |
| Stroke | At least 1 inpatient claim OR 2 HOP/Carrier claims with DX codes | ICD9: 430, 431, 433.01, 433.11, 433.21, 433.31, 433.81, 433.91, 434.00, 434.01, 434.10, 434.11, 434.90, 434.91, 435.0, 435.1, 435.3, 435.8, 435.9, 436, 997.02 (any DX on the claim) EXCLUSION: If any of the qualifying claims have: 800 <= DX Code <= 804.99, 850 <= DX Code <= 854.19 in any DX position OR DX V57xx as the principal DX Code, then EXCLUDE.  ICD10: G45.0, G45.1, G45.2, G45.8, G45.9, G46.0, G46.1, G46.2, G46.3, G46.4, G46.5, G46.6, G46.7, G46.8, G97.31, G97.32, I60.00, I60.01, I60.02, I60.10, I60.11, I60.12, I60.20, I60.21, I60.22, I60.30, I60.31, I60.32, I60.4, I60.50, I60.51, I60.52, I60.6, I60.7, I60.8, I60.9, I61.0, I61.1, I61.2, I61.3, I61.4, I61.5, I61.6, I61.8, I61.9, I63.00, I63.011, I63.012, I63.013, I63.019, I63.02, I63.031, I63.032, I63.039, I63.09, I63.10, I63.111, I63.112, I63.113, I63.119, I63.12, I63.131, I63.132, I63.133, I63.139, I63.19, I63.20, I63.211, I63.212, I63.213, I63.219, I63.22, I63.231, I63.232, I63.233, I63.239, I63.29, I63.30, I63.311, I63.312, I63.313, I63.319, I63.321, I63.322, I63.323, I63.329, I63.331, I63.332, I63.333, I63.339, I63.341, I63.342, I63.343, I63.349, I63.39, I63.40, I63.411, I63.412, I63.413, I63.419, I63.421, I63.422, I63.423, I63.429, I63.431, I63.432, I63.433, I63.439, I63.441, I63.442, I63.443, I63.449, I63.49, I63.50, I63.511, I63.512, I63.513, I63.519, I63.521, I63.522, I63.523, I63.529, I63.531, I63.532, I63.533, I63.539, I63.541, I63.542, I63.543, I63.549, I63.59, I63.6, I63.8, I63.81, I63.89, I63.9, I66.01, I66.02, I66.03, I66.09, I66.11, I66.12, I66.13, I66.19, I66.21, I66.22, I66.23, I66.29, I66.3, I66.8, I66.9, I67.841, I67.848, I67.89, I97.810, I97.811, I97.820, I97.821 (any DX on the claim) EXCLUSION: If any of the qualifying claims have any of the following codes in any DX position then EXCLUDE: S01.90XA, S02.0XXA, S02.0XXB, S02.101A, S02.101B, S02.102A, S02.102B, S02.109A, S02.109B, S02.10XA, S02.10XB, S02.110A, S02.110B, S02.111A, S02.111B, S02.112A, S02.112B, S02.113A, S02.113B, S02.118A, S02.118B, S02.119A, S02.119B, S02.11GA, S02.11GB, S02.11HA, S02.11HB, S02.121A, S02.121B, S02.121D, S02.121G, S02.121K, S02.121S, S02.122A, S02.122B, S02.122D, S02.122G, S02.122K, S02.122S, S02.129A, S02.129B, S02.129D, S02.129G, S02.129K, S02.129S, S02.19XA, S02.19XB, S02.2XXA, S02.2XXB, S02.30XA, S02.30XB, S02.31XA, S02.31XB, S02.32XA, S02.32XB, S02.3XXA, S02.3XXB, S02.400A, S02.400B, S02.401A, S02.401B, S02.402A, S02.402B, S02.40AA, S02.40AB, S02.40BA, S02.40BB, S02.40CA, S02.40CB, S02.40DA, S02.40DB, S02.40EA, S02.40EB, S02.40FA, S02.40FB, S02.411A, S02.411B, S02.412A, S02.412B, S02.413A, S02.413B, S02.42XA, S02.42XB, S02.600A, S02.600B, S02.601A, S02.601B, S02.602A, S02.602B, S02.609A, S02.609B, S02.610A, S02.610B, S02.611A, S02.611B, S02.612A, S02.612B, S02.61XA, S02.61XB, S02.620A, S02.620B, S02.621A, S02.621B, S02.622A, S02.622B, S02.62XA, S02.62XB, S02.630A, S02.630B, S02.631A, S02.631B, S02.632A, S02.632B, S02.63XA, S02.63XB, S02.640A, S02.640B, S02.641A, S02.641B, S02.642A, S02.642B, S02.64XA, S02.64XB, S02.650A, S02.650B, S02.651A, S02.651B, S02.652A, S02.652B, S02.65XA, S02.65XB, S02.66XA, S02.66XB, S02.670A, S02.670B, S02.671A, S02.671B, S02.672A, S02.672B, S02.67XA, S02.67XB, S02.69XA, S02.69XB, S02.80XA, S02.80XB, S02.81XA, S02.81XB, S02.82XA, S02.82XB, S02.831A, S02.831B, S02.831D, S02.831G, S02.831K, S02.831S, S02.832A, S02.832B, S02.832D, S02.832G, S02.832K, S02.832S, S02.839A, S02.839B, S02.839D, S02.839G, S02.839K, S02.839S, S02.841A, S02.841B, S02.841D, S02.841G, S02.841K, S02.841S, S02.842A, S02.842B, S02.842D, S02.842G, S02.842K, S02.842S, S02.849A, S02.849B, S02.849D, S02.849G, S02.849K, S02.849S, S02.85XA, S02.85XB, S02.85XD, S02.85XG, S02.85XK, S02.85XS, S02.8XXA, S02.8XXB, S02.91XA, S02.91XB, S02.92XA, S02.92XB, S06.0X0A, S06.0X1A, S06.0X2A, S06.0X3A, S06.0X4A, S06.0X5A, S06.0X6A, S06.0X7A, S06.0X8A, S06.0X9A, S06.1X0A, S06.1X1A, S06.1X2A, S06.1X3A, S06.1X4A, S06.1X5A, S06.1X6A, S06.1X7A, S06.1X8A, S06.1X9A, S06.2X0A, S06.2X1A, S06.2X2A, S06.2X3A, S06.2X4A, S06.2X5A, S06.2X6A, S06.2X7A, S06.2X8A, S06.2X9A, S06.300A, S06.301A, S06.302A, S06.303A, S06.304A, S06.305A, S06.306A, S06.307A, S06.308A, S06.309A, S06.310A, S06.311A, S06.312A, S06.313A, S06.314A, S06.315A, S06.316A, S06.317A, S06.318A, S06.319A, S06.320A, S06.321A, S06.322A, S06.323A, S06.324A, S06.325A, S06.326A, S06.327A, S06.328A, S06.329A, S06.330A, S06.331A, S06.332A, S06.333A, S06.334A, S06.335A, S06.336A, S06.337A, S06.338A, S06.339A, S06.340A, S06.341A, S06.342A, S06.343A, S06.344A, S06.345A, S06.346A, S06.347A, S06.348A, S06.349A, S06.350A, S06.351A, S06.352A, S06.353A, S06.354A, S06.355A, S06.356A, S06.357A, S06.358A, S06.359A, S06.360A, S06.361A, S06.362A, S06.363A, S06.364A, S06.365A, S06.366A, S06.367A, S06.368A, S06.369A, S06.370A, S06.371A, S06.372A, S06.373A, S06.374A, S06.375A, S06.376A, S06.377A, S06.378A, S06.379A, S06.380A, S06.381A, S06.382A, S06.383A, S06.384A, S06.385A, S06.386A, S06.387A, S06.388A, S06.389A, S06.4X0A, S06.4X1A, S06.4X2A, S06.4X3A, S06.4X4A, S06.4X5A, S06.4X6A, S06.4X7A, S06.4X8A, S06.4X9A, S06.5X0A, S06.5X1A, S06.5X2A, S06.5X3A, S06.5X4A, S06.5X5A, S06.5X6A, S06.5X7A, S06.5X8A, S06.5X9A, S06.6X0A, S06.6X1A, S06.6X2A, S06.6X3A, S06.6X4A, S06.6X5A, S06.6X6A, S06.6X7A, S06.6X8A, S06.6X9A, S06.810A, S06.811A, S06.812A, S06.813A, S06.814A, S06.815A, S06.816A, S06.817A, S06.818A, S06.819A, S06.820A, S06.821A, S06.822A, S06.823A, S06.824A, S06.825A, S06.826A, S06.827A, S06.828A, S06.829A, S06.890A, S06.891A, S06.892A, S06.893A, S06.894A, S06.895A, S06.896A, S06.897A, S06.898A, S06.899A, S06.9X0A, S06.9X1A, S06.9X2A, S06.9X3A, S06.9X4A, S06.9X5A, S06.9X6A, S06.9X7A, S06.9X8A, S06.9X9A, OR Z51.89 as the principal DX Code then EXCLUDE. |
| Anticoagulant prescription | Medicare part D prescription data | Prescription for any of the following medications during follow-up: warfarin, apixaban, rivaroxaban, edoxaban, betrixaban, dabigatran, argatroban, hirudin, bivalirudin, desirudin, enoxaparin, dalteparin, fondaparinux, tinzaparin |
